# Role of Drugs used for chronic disease management on Susceptibility and Severity of COVID-19: A Large Case-Control Study

**DOI:** 10.1101/2020.04.24.20077875

**Authors:** Huadong Yan, Ana M Valdes, Amrita Vijay, Shanbo Wang, Lili Liang, Shiqing Yang, Hongxia Wang, Xiaoyan Tan, Jingyuan Du, Susu Jin, Kecheng Huang, Fanrong Jiang, Shun Zhang, Nanhong Zheng, Yaoren Hu, Ting Cai, Guruprasad P. Aithal

**Affiliations:** Department of Infectious Diseases, Hwamei Hospital, Ningbo No.2 Hospital, University of Chinese Academy of Sciences., Ningbo 315010, China; Department of New Medical Science, Key Laboratory of Diagnosis and Treatment of Digestive System Tumors of Zhejiang Province, Hwamei Hospital, Ningbo No.2 Hospital, University of Chinese Academy of Sciences, Ningbo 315010, China; NIHR Nottingham Biomedical Research Centre, Nottingham University Hospitals NHS Trust and University of Nottingham, Nottingham, UK; Division of Rheumatology, Orthopaedics and Dermatology, School of Medicine, University of Nottingham, Nottingham; Nottingham Digestive Diseases Centre, School of Medicine, University of Nottingham, Nottingham UK; Department of General Practice, Gulin Township Health Center, Ningbo 315010, China; Department of Infectious Diseases, Ninghai No.1 Hospital, Ningbo 315600, China; Department of Pharmacology, Hwamei Hospital, Ningbo No.2 Hospital, University of Chinese Academy of Sciences., Ningbo 315010, China

**Author notes:** Joint first authors (Huadong Yan, Ana M Valdes). Joint Corresponding authors: Guruprasad P. Aithal, Nottingham Digestive Diseases Centre, School of Medicine, University of Nottingham, Nottingham UK. Email address. Amrita Vijay, Nottingham Digestive Diseases Centre, School of Medicine, University of Nottingham, Nottingham UK. Email address. Ting Cai, Department of Infectious Diseases, Key Laboratory of Diagnosis and Treatment of Digestive System Tumors of Zhejiang Province, Hwamei Hospital, Ningbo No.2 Hospital, University of Chinese Academy of Sciences, Ningbo 315010, China.

**Keywords:** COVID-19, Antihypertensive medications, Antidiabetic medications, Corticosteroids, Calcium channel blockers, Severity, Risk

## Abstract

The study aimed to investigate whether specific medications used in the treatment chronic diseases affected either the development and/ or severity of COVID-19 in a cohort of 610 COVID-19 cases and 48,667 population-based controls from Zheijang, China. Using a cohort of 578 COVID-19 cases and 48,667 population-based controls from Zheijang, China we tested the role of usage of cardiovascular, antidiabetic and other medications on risk and severity of COVID 19. Analyses were adjusted for age, sex and BMI and for presence of relevant comorbidities. Individuals with hypertension taking calcium channel blockers had significantly increased risk [odds ratio (OR)= 1.73 (95% CI 1.2-2.3)] of manifesting symptoms of COVID-19 whereas those taking angiotensin receptor blockers and diuretics had significantly lower disease risk (OR=0.22; 95%CI 0.15-0.30 and OR=0.30; 95%CI 0.19-0.58 respectively). Among those with type 2 diabetes, dipeptidyl peptidase-4 inhibitors (OR= 6.02; 95% CI 2.3-15.5) and insulin (OR= 2.71; 95% CI 1.6-5.5) were more and glucosidase inhibitors were less prevalent (OR= 0.11; 95% CI 0.1-0.3) among with COVID-19 patients.

Drugs used in the treatment of hypertension and diabetes influence the risk of development of COVID-19, but, not its severity.

**Study highlights:** *What is the current knowledge on the topic?:* Cardiovascular disease and Diabetes have been highlighted as comorbidities contributing to a more severe form of COVID-19 and medication to treat them may also influence the risk of COVID-19 and its clinical outcomes.

*What question did this study address?:* Does specific medications used in the treatment of chronic diseases influence the risk for the susceptibility to SARS CoV-2 infection of severity of COVID-19?

*What does this study add to our knowledge?:* The study confirms that higher BMI, diabetes and cardio/ cerebrovascular disease as independent risk factors for the development of COVID-19. Angtiotensin Receptor Blockers (ARBs) and diuretics were associated with reduced risk and Calcium Channel Blockers (CCBs) with increased risk of developing COVID-19. Among those with type 2 diabetes, dipeptidyl peptidase-4 and were associated with increased and glucosidase inhibitors with reduced risk development of COVID-19. None of the antihypertensive or anti-diabetic drugs were associated with increased risk of severe or critical form of the infection. Drugs used in the treatment of hypertension and diabetes influence the risk of development of COVID-19, but are not associated with severity of the disease.

*How might this change clinical pharmacology or translational science?:* Findings from the current large case-control study confirmed no evidence to alter ARBs or ACEIs therapy in the context of COVID-19 severity in clinical practice. Hypertension significantly increases the risk of severe or critical SARS-CoV-2 infection indicating that carefully controlled blood pressure should be a priority to reduce the healthcare burden of COVID-19.

## Introduction

The pandemic of coronavirus disease 2019 (COVID-19) caused by a new zoonotic coronary virus, SARS CoV2, has affected over 8.5 million people and caused over 450,000 deaths across the world as of 21 June 2020^1^ having a profound impact on health services and public health. The clinical spectrum of SARS-CoV-2 infection appears to be wide, encompassing asymptomatic infection in a minimum of 5%, mild upper respiratory tract illness with fever, fatigue, cough with or without sputum production in 81%, to more severe viral pneumonia in 14% with ground-glass opacity computed tomography of the chest leading to critical illness in 5% associated with respiratory failure, septic shock and/ or multi-organ failure.^2^ Case-fatality rate ranges from 0.4% to 2.9% on average between different regions.^2^

Large descriptive studies have reported type 2 diabetes in 7.4% - 9.6% of patients with COVID-19 and hypertension in 15% as two of the most prevalent co-morbid conditions. ^3,4,5^ Moreover, diabetes (16.2%) and hypertension (23.7%) are highly prevalent among those with severe manifestations.^3^ Of patients admitted to intensive care unit, 22% of those who died had diabetes.^6^

SARS CoV2 virus binds angiotensin-converting enzyme 2 (ACE2); ^7,8^ in humans, ACE2 is expressed broadly including epithelial cells of the lung, intestine, kidney, heart and blood vessels.^9^ The virus downregulates the ACE2 protein expression in a replication dependent manner^10^ resulting in loss of ACE2 function. Whether variation in ACE2 expression contributes to the virulence in the current pandemic of COVID-19 is still unclear. There is an ongoing debate as to whether and how the interaction between the virus, hypertension and ACE2 may influence the manifestations of COVID-19.^11,12,13^ Concerns have been raised that angiotensin converting enzyme inhibitors (ACEIs), angiotensin II type 1 receptor blockers (ARBs) and thiozolidinediones often prescribed in patients with diabetes, hypertension and cardiac disease may increase the risk of COVID-19 and its clinical outcomes.^11,12,14^ In contrast, ACE2 is a counter regulatory enzyme that degrades angiotensin II, hence, reducing its effect on vasoconstriction, sodium retention and fibrosis. Therefore, trials of losartan as a treatment for Covid-19 are underway enrolling patients who have not previously received treatment with ACEIs or ARBs.^15^ In addition, dipeptidyl peptidase-4 (DPP4), the receptor for Middle East respiratory syndrome-related coronavirus (MERS-CoV) has been shown to have similar expression profile to ACE2 in the lung.^16^ DPP4 inhibitors are commonly used in the treatment of type 2 diabetes.

We have investigated whether any of the medications for chronic conditions, in particular antihypertensive and antidiabetic medications affected either the development and/ or severity of COVID-19 using a multi-center cohort of 578 patients with common, severe or critical form of COVID-19 and 48,667 population-based control subjects from Zhejiang province, China.

## Patients and Methods

The study protocol conformed to the ethical guidelines of the 1975 Declaration of Helsinki. The local ethics committees of all hospitals approved the retrospective study of cohorts COVID-19. The requirement for written consent was waived due to the retrospective and anonymous nature of this study.

### Cohort of Cases

Consecutive patients presenting to 14 hospitals in Zhejiang province, China (see collaborators) between Jan 10 and Feb 28, 2020 and confirmed diagnosis of COVID-19 infection were included (**Figure 1**). The diagnosis of COVID-19 was made in accordance with the Guidelines for the Diagnosis and Treatment of New Coronavirus Pneumonia (fifth edition) formulated by the National Health Commission of the People’s Republic of China.^17^ confirmed case of COVID-19 was defined as a positive result on real-time reverse-transcriptase–polymerase-chain-reaction (RT-PCR) assay of nasal and pharyngeal swab specimens. Only laboratory-confirmed cases were included in the study. The clinical data of all patients were collected from the electronic medical records. All patients were administered with antiviral and supportive treatment, and prevention of complications based on their clinical condition.

**Figure 1:**
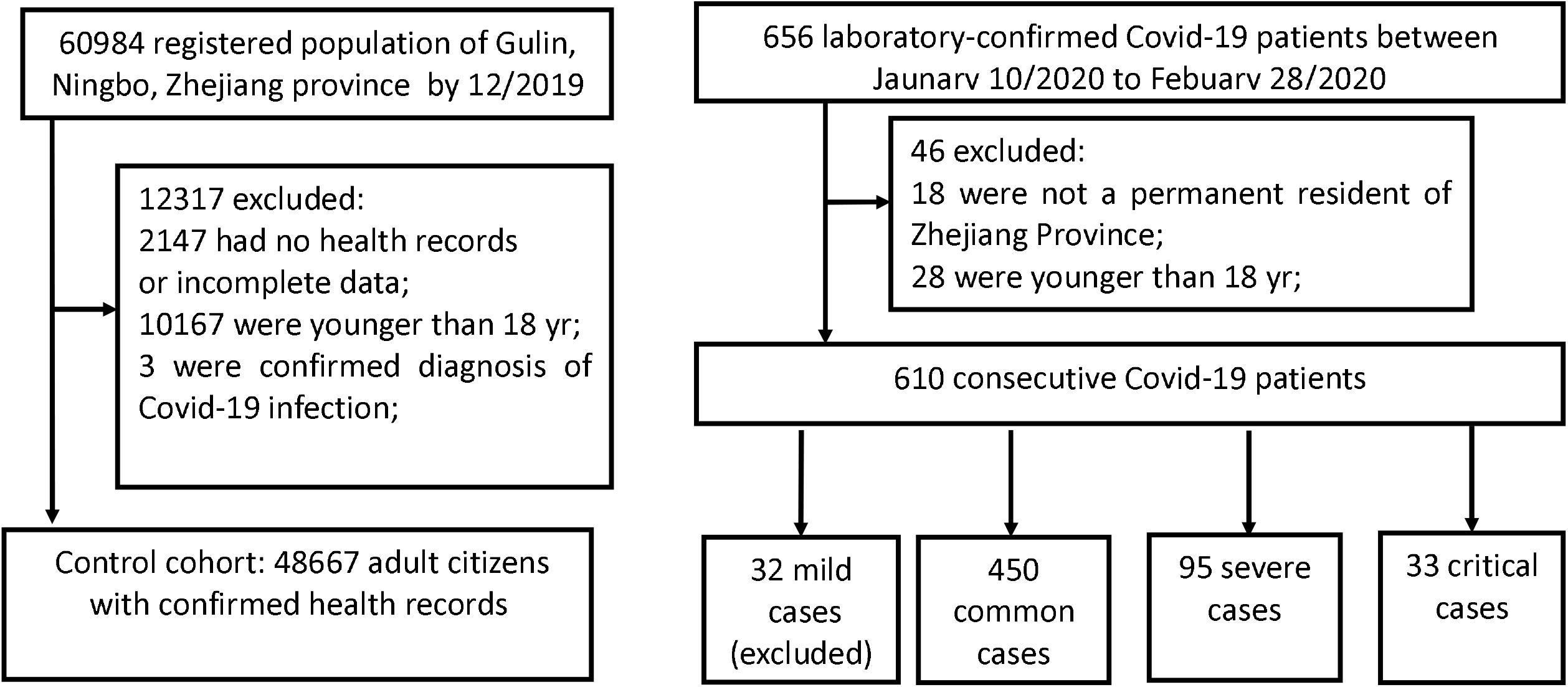
Flow chart of recruitment paths for research study participants

### Clinical categories

The severity of the disease was classified into 4 categories according to the Guidelines for the Diagnosis and Treatment of New Coronavirus Pneumonia (fifth edition)^17^: 1) *mild type*: patients with mild clinical symptoms and no pulmonary changes on CT imaging, 2) *common type*: patients with symptoms of fever and signs of respiratory infection, and having pneumonia changes on CT imaging, 3) *severe type*: patients presenting with any one item of the following: (a) respiratory distress, respiratory rate ≥ 30/min, (b) oxygen saturation of finger ≤ 93% in resting condition, and (c) arterial partial pressure of oxygen (PaO2) /oxygen concentration (FiO2) ≤ 300 mmHg (1 mmHg = 0.133 kPa) or 4) *critical type*: patients meeting any one of the following criteria: (a) respiratory failure requiring mechanical ventilation, (b) shock and (c) requiring ICU admission requirement due to multiple organ failure. Since only 32 mild cases were available and they would normally not be hospitalized, these were removed from the analysis (Figure 1).

### Population-based Control group

Detailed methods and data sources used to establish an EHR-based general population-based cohort study in an eastern coastal area of China has been described previously.^18^ The control group used in the current study is from the general population from Gulin town, an area of Ningbo City, Zhejiang province, China which covers the catchment area one of the hospitals from which one quarter of the COVID-19 patients for this study were recruited. Since 2004, all citizens are offered regular health screening in Gulin Health Center. Detailed medical history is recorded in Electronic Medical Record System. Gulin town was selected as a centre of the ‘Mega-Project for National Science and Technology Development’ under the “13th Five-Year Plan of China” for liver diseases between 2017 and January 2021. We included all adult citizens had detailed medical records established by 2019. A total of 48,667 adults from 60984 registered population who had electronic medical records were included in this study (**Figure 1**). The following data were collected: 1) age, 2) sex, 3) detailed medical history including diabetes, hypertension, malignancy, other severe diseases (congestive heart failure, severe neurological diseases and previous organ transplantation), 4) details of prescription drugs used to treat these medical conditions and 5) personal health parameters including history of alcohol consumption and smoking as well as anthropometric measurements including body weight, height, waist circumference, and arterial pressure as well as 6) laboratory parameters including serology for hepatitis B and C (HBsAg and anti-HCV). Individuals in the control cohort whose health record showed a diagnosis of COVID-19 (n=3) were removed from the analysis (Figure 1).

Health information systems in Gulin include different administrative databases of general demographic characteristics, health check information, health insurance database, inpatient and outpatient electronic medical records, chronic disease management (diabetes, hypertension, malignancy, myocardial Infarction) and death certificates, and so on. These databases are inherently linked to each other by a unique and encoded identifier for each individual. The system was originally designed in 2006 to facilitate routine primary care services for local general practitioners (GPs). Since 2009, this regional system has covered nearly all health-related activities of residents within this region, from birth to death, including children, adolescents, pregnant women, adults and elderly people. Now 98% of permanent residents are covered by the national health insurance and have registered in the health information system with a valid healthcare identifier.

All the chronic diseases diagnosed by doctors in Gulin or outside Gulin will be reported to Ningbo CDC and rechecked by department of chronic disease management of Gulin hospital. All the prescription medicines of chronic diseases listed above are recorded and regularly verified.

*Use of medication:* it is the role of Dept. of Chronic Diseases within each community hospital in Zheijang province to update the prescriptions of chronic diseases such as diabetes, hypertension, cardiovascular, cerebrovascular disease and tumors regularly. From these data we were able to run a database search including for this study and individuals were classified as taking a given medication if they had been given a prescription for a that drug at least three times. For COVID-19 patients, the medication was included only if 3 or more one-month prescriptions were filled <u>before</u> diagnosis of COVID-19, according to the patient’s records.

### Statistical methods

Categorical variables were expressed as frequency and percentages. Continuous variables were expressed as mean and standard deviations. Logistic regressions were carried out either unadjusted or adjusting for age, sex and BMI. P<0.05 was considered statistically significant.

## Results

The selection of the study population is illustrated in **Figure 1**. A total of 610 COVID-19 patients were enrolled after admission from various centers in the Zheijiang province (**Figure 1**). A population-based cohort from the Zhejiang province consisting of 48,667 adults with health records was used as controls. The descriptive characteristics, comorbidities and use of medication prior to admission available in both cases and controls were compared (**Table 1**). We found that, prevalence of type 2 diabetes, higher BMI, presence of cardiovascular or cerebrovascular disease were associated with (all increased in) the covid-19 patients (**Table 1**). We then adjusted all factors for age, sex and BMI. In the case of antihypertensives we further adjusted for hypertension, in the case of medication used to treat diabetes we adjusted for presence of T2D.

**Table 1.** Comparison between Covid-19 cases and controls, showing univariate odds ratios from logistic regression, and odds ratios adjusted for age, sex and BMI

|  |  |  |  | unadjusted |  |  |  | adjusted for age, sex, BMI |  |  |
| --- | --- | --- | --- | --- | --- | --- | --- | --- | --- | --- |
|  | controls | COVID-19 | OR | 95% CI |  | p-value | OR | 95% CI |  | p-value |
| **n** | 48667 | 578 |  |  |  |  |  |  |  |  |
| **sex M** | 23506 | 293 | 1.10 | 0.93 | 1.29 | 0.25 |  |  |  |  |
| **n (%)** | 48.3% | (50.7%) |  |  |  |  |  |  |  |  |
| **Age years mean (SD)** | 49.96 (16.69) | 49.18 (14.16) | 1.00 | 0.99 | 1.00 | 0.273 |  |  |  |  |
| **BMI kg/m<sup>2</sup> mean (SD)** | 22.79 (2.86) | 24.01 (3.43) | 1.13 | 1.10 | 1.16 | <0.0001 |  |  |  |  |
| **Smoke** | 4725 | 53 | 0.90 | 0.66 | 1.19 | 0.477 | 0.71 | 0.51 | 0.98 | 0.071 |
| **n (%)** | (9.7%) | (9.2%) |  |  |  |  |  |  |  |  |
| **HBV** | 2287 | 32 | 1.22 | 0.84 | 1.71 | 0.275 | 1.27 | 0.86 | 1.81 | 0.201 |
| **n (%)** | (4.7%) | (5.5%) |  |  |  |  |  |  |  |  |
| **Diabetes** | 2920 | 57 | 1.70 | 1.28 | 2.22 | 0.0001 | 1.53 | 1.15 | 2.11 | 0.004 |
| **n (%)** | (6.1%) | (9.8%) |  |  |  |  |  |  |  |  |
| **Hypertension** | 9855 | 130 | 1.18 | 0.97 | 1.43 | 0.084 | 1.01 | 0.76 | 1.31 | 0.851 |
| **n (%)** | (20.2%) | (22.4%) |  |  |  |  |  |  |  |  |
| **Cardio or cerebrovascular disease n (%)** | 622<br>(1.3%) | 15<br>(2.6%) | 2.16 | 1.25 | 3.45 | 0.002 | 2.26 | 1.22 | 3.76 | 0.006 |
| **Tumour** | 973 | 10 | 0.86 | 0.43 | 1.53 | 0.648 | 0.92 | 0.51 | 1.73 | 0.991 |
| **n (%)** | (2.0%) | (1.8%) |  |  |  |  |  |  |  |  |
| **Glucocorticoids** | 326 | 5 | 1.29 | 0.46 | 2.82 | 0.568 | 1.17 | 0.41 | 2.73 | 0.717 |
| **n (%)** | (0.6%) | (0.8%) |  |  |  |  |  |  |  |  |
| **ACEI** | 555 | 5 | 0.75 | 0.27 | 1.64 | 0.538 | 0.61<sup>(1)</sup> | 0.23 | 1.51 | 0.362 |
| **n (%)** | (1.1%) | (0.8%) |  |  |  |  |  |  |  |  |
| **ARB** | 7485 | 50 | 0.56 | 0.41 | 0.73 | <0.0001 | 0.22<sup>(1)</sup> | 0.15 | 0.30 | <0.0001 |
| **n (%)** | (15.4%) | (8.7%) |  |  |  |  |  |  |  |  |
| **CCB** | 4740 | 84 | 1.61 | 1.27 | 2.01 | <0.0001 | 1.73<sup>(1)</sup> | 1.21 | 2.31 | 0.001 |
| **n (%)** | (9.7%) | (14.6%) |  |  |  |  |  |  |  |  |
| **Diuretic** | 2467 | 12.31 | 0.43 | 0.24 | 0.72 | 0.003 | 0.30<sup>(1)</sup> | 0.19 | 0.58 | <0.0001 |
| **n (%)** | (5.1%) | (2.1%) |  |  |  |  |  |  |  |  |
| **Beta.blockers** | 1221.54 | 15 | 0.96 | 0.53 | 1.57 | 0.892 | 0.80<sup>(1)</sup> | 0.42 | 1.47 | 0.468 |
| **n (%)** | (2.5%) | (2.3%) |  |  |  |  |  |  |  |  |
| **Glycosidase inhibitors** | 2019 | 15 | 0.66 | 0.38 | 1.05 | 0.104 | 0.11<sup>(2)</sup> | 0.10 | 0.41 | <0.0001 |
| **n (%)** | (4.1%) | (2.6%) |  |  |  |  |  |  |  |  |
| **Biguanides** | 1445 | 28 | 1.64 | 1.09 | 2.37 | 0.010 | 1.00<sup>(2)</sup> | 0.58 | 1.62 | 0.975 |
| **n (%)** | (2.9%) | (4.9%) |  |  |  |  |  |  |  |  |
| **Insulin secretagogues** | 700 | 16 | 1.92 | 1.12 | 3.06 | 0.010 | 1.23<sup>(2)</sup> | 0.72 | 2.29 | 0.395 |
| **n (%)** | (1.4%) | (2.8%) |  |  |  |  |  |  |  |  |
| **Thiazolidinediones** | 394 | 3 | 0.64 | 0.16 | 1.68 | 0.446 | 0.36<sup>(2)</sup> | 0.11 | 1.21 | 0.121 |
| **n (%)** | (0.8%) | (0.5%) |  |  |  |  |  |  |  |  |
| **DPP-4 inhibitors** | 486 | 6 | 9.22 | 3.54 | 19.82 | <0.0001 | 6.02<sup>(2)</sup> | 2.30 | 15.51 | <0.0001 |
| **n (%)** | (0.1%) | (0.9%) |  |  |  |  |  |  |  |  |
| **Insulin** | 701 | 16 | 3.29 | 1.86 | 5.34 | <0.0001 | 2.71<sup>(2)</sup> | 1.57 | 5.51 | 0.0002 |
| **n (%)** | (1.4%) | (2.8%) |  |  |  |  |  |  |  |  |
| **Statins** | 443 | 15 | 2.82 | 1.60 | 4.57 | <0.0001 | 2.50 | 1.48 | 4.21 | 0.001 |
| **n (%)** | (0.9%) | (2.6%) |  |  |  |  |  |  |  |  |
| **Fibrates** | 63 | 0 | 0.05 | 0.00 | 30.12 | 0.417 | 0.03 | 0.00 | 29.26 | 0.339 |
| **n (%)** | (0.1%) | (0.0%) |  |  |  |  |  |  |  |  |
| **Aspirin** | 673 | 10 | 1.45 | 0.75 | 2.53 | 0.220 | 1.28 | 0.57 | 2.63 | 0.512 |
| **n (%)** | (1.3%) | (1.8%) |  |  |  |  |  |  |  |  |

<sup>(1)</sup> in n=134 covid-19 patients with hypertension
<sup>(2)</sup> in n=58 covid-19 patients with Type 2 Diabetes

All the disease conditions associated with COVID-19 remain statistically significantly associated after adjustment for age, sex and BMI. There was no significant difference in the use of glucocorticoids between COVID-19 and the population-based controls and a significantly higher use of statins among COVID-19.

We found that among individuals with a diagnosis of hypertension there was no difference in the use of ACEIs or beta-blockers, but individuals with COVID-19 were significantly more likely to be on calcium channel blockers (CCBs) and less likely to be on diuretics and on ARBs.

We also found that after adjustment for demographics and presence of T2D the use of antidiabetic medications was significantly different in T2D patients with COVID-19 than in those without and this is particularly striking for use of glycosidase inhibitors which are significantly less prevalent among T2D individuals infected with COVID-19, and insulin and DPP-4 inhibitors which are much more likely to be used by T2D people with COVID-19. (**Table 1**)

We compared these figures graphically between controls and different severity categories in **Figure 2**. We see that although hypertension in itself is not more prevalent in COVID-19 diagnosed individuals than controls, it is much more prevalent in severe and critical cases (**Table 2**, **Figure 2A**). Type 2 diabetes, male sex, and age over 65 all show a similar pattern in terms of increase prevalence with severity. (**Table 2**, **Figure 2A**)

**Figure 2.**
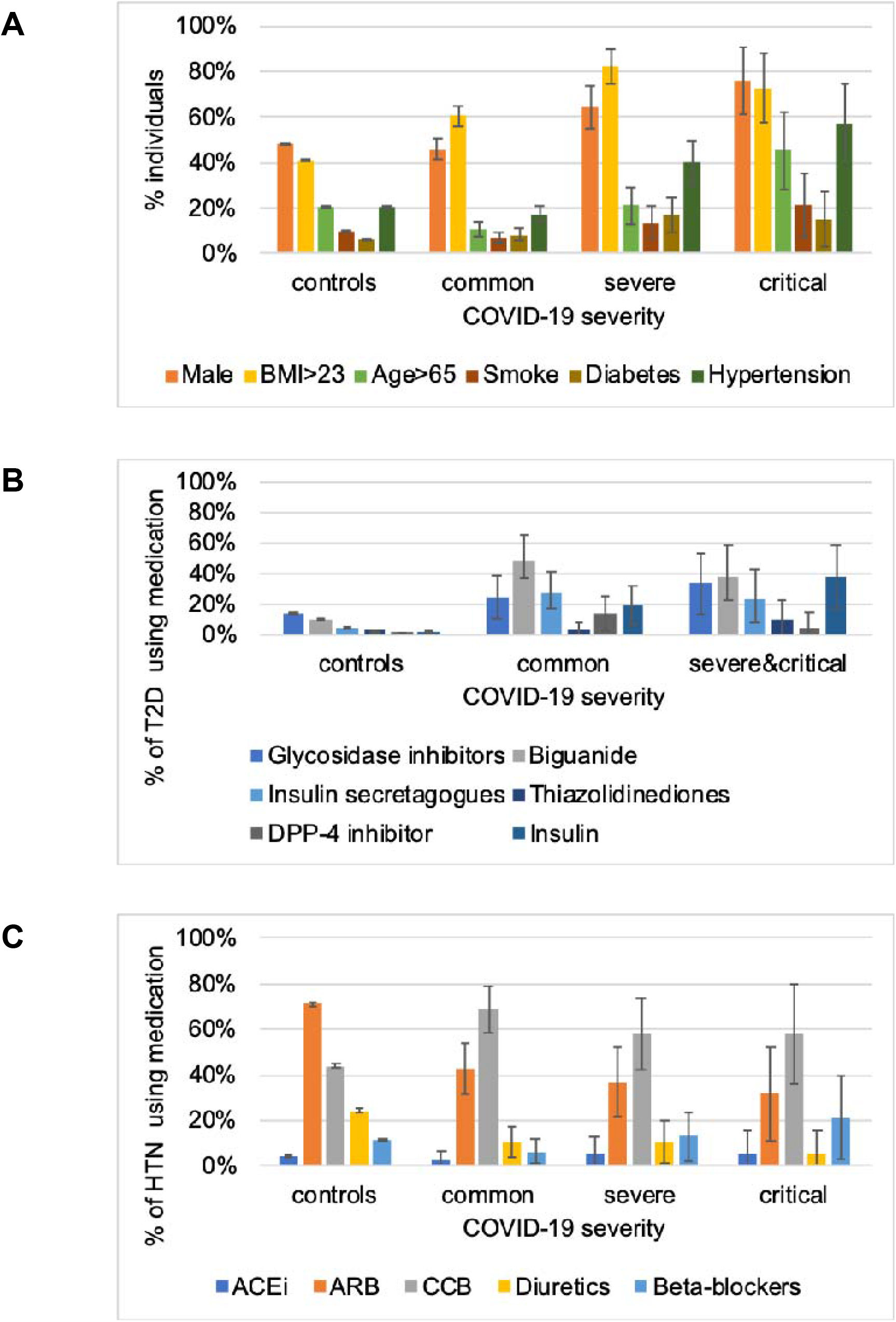
**(A)** Comparison of demographic and co-morbidity distribution between COVID-19 positive individuals grouped according to severity compared to the control cohort. **(B)** Comparison of use of type 2 diabetes (T2D) prescribed medications across the COVID-19 severity range compared to the control cohort. Error bars represent 95% CI. **(C)** Comparison of use of anti hypertensive (HTN) prescribed medications across the COVID-19 severity range compared to the control cohort. Error bars represent 95% CI.

**Table 2.** Association between demographics, clinical presentation at admission, comorbidities and use of medication and COVID-19 severity. odds ratios were derived by logistic regression severe (severe+ critical) vs not severe (common). p-values adjusted for age, sex and BMI, except those for age, sex and BMI

| Severity category | 2<br>(common) | 3+4<br>(severe + critical) | adj OR<br>severe vs not | 95% CI |  | p-value |
| --- | --- | --- | --- | --- | --- | --- |
| n | 450 | 128 |  |  |  |  |
| sex M (n, %) | 207,<br>46% | 86,<br>67% | 2.40 | 1.59 | 3.65 | P<0.0001 |
| Age years<br>mean (SD) | 47.26<br>(13.53) | 56<br>(14.29) | 1.04 | 1.03 | 1.06 | P<0.0001 |
| BMI kg/m2<br>mean (SD) | 23.63<br>(3.29) | 25.38<br>(3.59) | 1.15 | 1.09 | 1.23 | P<0.0001 |
| *Clinical presentation* |  |  |  |  |  |  |
| Fever<br>n (%) | 365.5<br>(81.3%) | 109.9<br>(85.9%) | 1.61 | 0.42 | 2.50 | 0.111 |
| Cough<br>n (%) | 296.64<br>(65.9%) | 102.98<br>(80.4%) | 2.13 | 1.33 | 3.49 | 0.002 |
| digestive symptoms<br>n (%) | 52.02<br>(11.5%) | 20<br>(15.6%) | 1.42 | 0.80 | 2.44 | 0.220 |
| systolic BP mmHg<br>mean (SD) | 131.65<br>(19.29) | 133<br>(18.17) | 0.97 | 0.95 | 1.30 | 0.011 |
| diastolic BP mmHg<br>mean (SD) | 81.07<br>(10.85) | 78.75<br>(10.08) | 0.96 | 0.93 | 0.99 | 0.009 |
| respiratory rate<br>mean (SD) | 19.42<br>(1.80) | 20.86<br>(4.58) | 1.22 | 1.08 | 1.33 | 0.002 |
| peak temperature C<br>mean (SD) | 37.88<br>(0.77) | 38.08<br>(0.85) | 1.38 | 1.06 | 1.80 | 0.018 |
| days since disease<br>onset<br>mean (SD) | 6.15<br>(5.15) | 6.57<br>(3.99) | 1.01 | 0.97 | 1.01 | 0.637 |
| hospital length of<br>stay<br>mean (SD) | 19.31<br>(18.63) | 21.18<br>(9.87) | 1.00 | 1.00 | 1.01 | 0.363 |
| Death<br>n (%) | 0<br>0.00% | 16.5<br>12.90% | 1054.07 | 0.00 | inf | 0.350 |
| **Comorbidities** |  |  |  |  |  |  |
| **Smoke**<br>**n (%)** | 31<br>(6.9%) | 20<br>(15.6%) | 0.88 | 0.45 | 1.74 | 0.728 |
| **Chronic Liver**<br>**Disease**<br>**n (%)** | 27<br>(6.0%) | 6<br>(4.6%) | 0.77 | 0.28 | 1.80 | 0.573 |
| **Diabetes**<br>**n (%)** | 37<br>8.2% | 21<br>15.6% | 1.13 | 0.64 | 1.41 | 0.570 |
| **Hypertension**<br>**n (%)** | 77<br>(17.1%) | 57<br>(44.5%) | 2.88 | 1.54 | 4.96 | 0.0007 |
| **CVD cerebrovascular**<br>**n (%)** | 10<br>2.2% | 6<br>4.6% | 1.12 | 0.37 | 3.39 | 0.841 |
| **Cancer**<br>**n (%)** | 6<br>1.3% | 4<br>3.1% | 1.38 | 0.42 | 7.18 | 0.284 |

With regards to antidiabetic medications after adjusting for age, sex, BMI and diabetes, glucosidase inhibitors are less prevalent among T2D affected individuals symptomatic for COVID-19 and DPP-inhibitors and insulin are much more prevalent among COVID-19 patients. We compared these values to those in COVID-19 severity categories (**Figure 2B** and **Table 3**) and we find that there is no significant association between use of DPP-inhibitors and COVID-19 severity after adjustment for age, sex and BMI, resulting in OR=0.32 (95%CI 0.02 -2.18; P=0.31). For insulin, we observed a trend for higher use among the severe cases with OR=2.63 (95%CI 0.80-9.07 P=0.11) which however did not achieve statistical significance (**Table 3**).

**Table 3.**
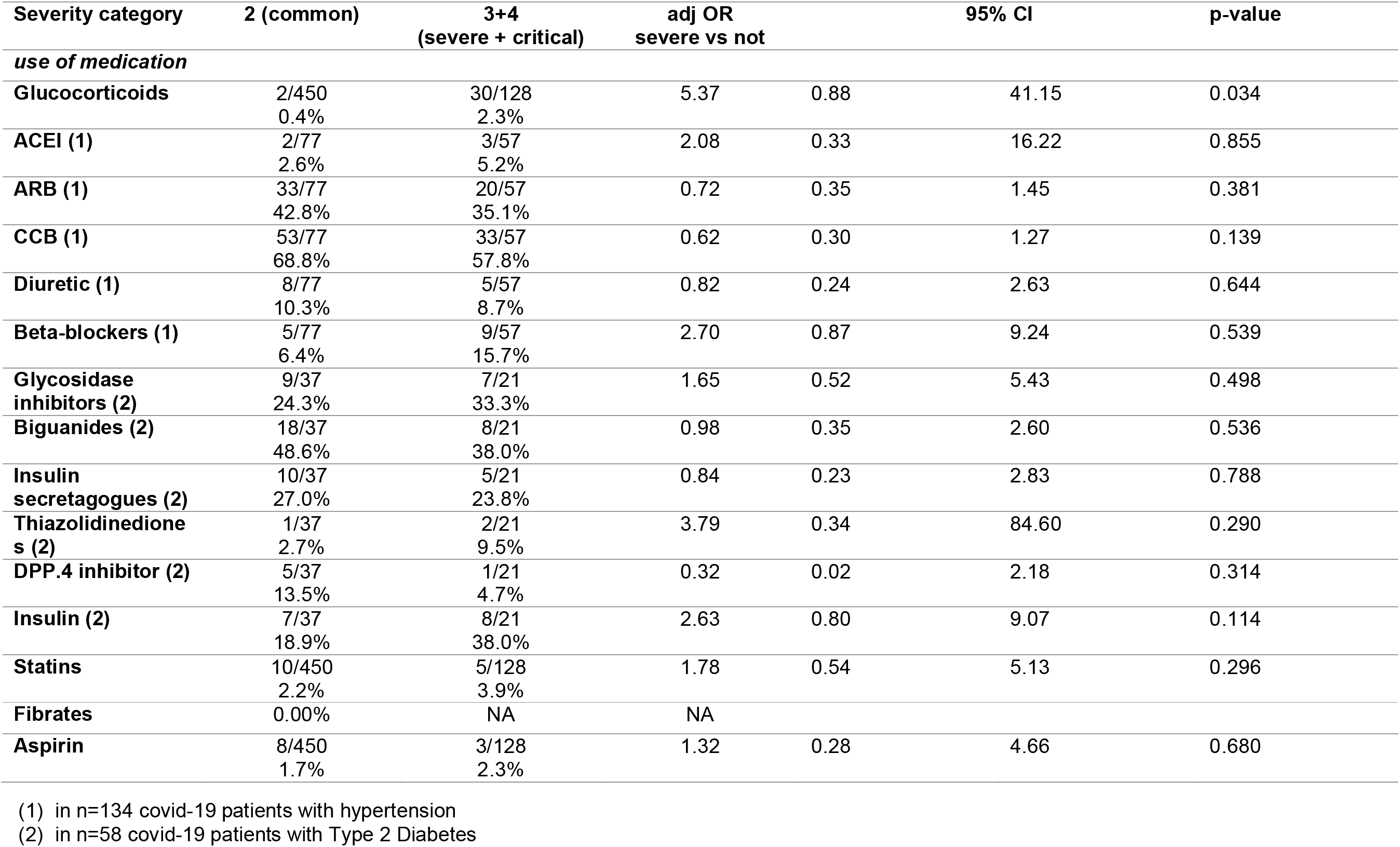
Association between the use of medication and COVID-19 severity. odds ratios were derived by logistic regression severe severe+ critical) vs not severe (common). p-values adjusted for age, sex and BMI

We carried out a similar analysis with antihypertensive medications. We found that comparing population-based controls to COVID-19 diagnosed individuals that use of ARBs and diuretics was significantly less prevalent in COVID-19 cases with hypertension than among hypertensive controls, whilst use of CCBs was significantly more prevalent among COVID-19 cases than controls after adjustment for age, sex and BMI (**Table 1**). No significant difference in susceptibility or severity was seen with use of ACEIs. (**Figure 2C**, **Table 3**). In terms of severity, there was a trend for use of beta-blockers to be more prevalent in severe and critical cases than in mild and common cases. However, after adjustment for age, sex and BMI none of the antihypertensive medications was significantly associated with COVID-19 severity. (**Figure 2C, Table 3**)

The only type of medications which remained associated with COVID-19 severity after adjustment for age, sex and BMI were immunosuppressive drugs (glucocorticoids) with an OR =5.37 (95%CI 0.88-41.15; **Table 3**)

## Discussion

Recent descriptive cohort studies have reported type 2 diabetes in 7.4% - 9.6% of patients with COVID-19 and hypertension in 15% as two of the most prevalent chronic health conditions.^3-5,19^ Using a case-control design involving 610 patients with COVID-19 and a well-characterized 48,667 population-based controls from the South-east region of China, we have identified higher BMI, diabetes and cardio/cerebrovascular disease as independent risk factors for the development of COVID-19. We found the prevalence of hypertension to be slightly higher among patients (22.4%), but this was not statistically significant compared to its prevalence of 20.2% in the population-based control group. Furthermore, age (>65 years), sex (male) and BMI were associated with the development of severe disease in 128 out of 610 of those with COVID-19. In addition, when adjusted for age, sex and BMI, those with history of hypertension have over two-fold risk of developing severe form of COVID-19.

The analysis of medications acting on renin-angiotensin-aldosterone system demonstrates that patients with COVID-19 were significantly less likely compared to population-based controls to be taking ARBs (8.7% vs 15.4%) and diuretics in (2.1% vs 5.1%) prior to their presentation, after adjusting for covariates such as age, sex and BMI) as well as presence of hypertension. In contrast, CCBs were associated with increased risk of COVID-19. Although hypertension is clearly a risk factor for severity, we found that none of the anti-hypertensive medications were positively associated with severity of the disease. In the case-control analysis, patients with type 2 diabetes, treatment with dipeptidyl peptidase-4 inhibitors and insulin were associated with increased risk of COVID-19 while glucosidase inhibitor therapy was associated with reduced risk. It has been suggested that DPP4 residues might interact with SARS-CoV-2 S1 domain of the Spike protein, also targeted by other coronaviruses that enter the host cells through the functional receptor DPP4.^20^ In addition to its catalytic functions DPP4 also has a role in immune mechanisms and inflammatory process^20^ which may be influenced by DPP4 inhibitors. Treatment with insulin may also reflect the degree of insulin resistance, a well-established risk factor for COVID-19. Alpha glucosidase inhibitors improve glycemic control by acting on intestinal membrane-bound enzymes; their effect on the cellular entry of SARS CoV-2^21^ is unclear. In contrast to their association with the development of COVID-19, none of the antidiabetic drugs were associated with the severe or critical form of the disease. To our knowledge, these associations of antidiabetic medications with COVID-19 infection haven’t been demonstrated so far. The higher intake of anti-diabetic medications might be linked to a longer duration or lower glycaemic control and points to the role of metabolic inflammation in predisposing individuals to developing the more severe form of the disease.^22^

Among medications, corticosteroids intake at baseline were the only group that increased the risk of severity of COVID-19, but this affected only 2.1% of patients. In the study from OpenSAFELY collaborative, patients with asthma with recent oral steroid treatment had 25% additional risk of in-hospital mortality. ^23^ Although in the latter study, oral steroid treatment was used as a marker of severity, the effect on the disease outcome could be related to recent exposure to corticosteroid.

The concerns raised regarding the safety of ACEIs and ARBs in relation to COVID-19, are based on the hypothesis that medications acting on renin-angiotensin-aldosterone system may raise the expression of ACE2, postulated receptor for SARS-CoV-2.^14,24-26^ The virus uses human ACE2 ^27^ expressed in lung alveolar epithelial cells to trigger the key manifestations and complications of the infection. ACE2 is involved in the hydrolysis of angiotensin I and II into inactive forms 1–9 and 1–7 respectively.^15^ Angiotensin 1–7 act on the Mas receptor to play a protective role through vasodilatory and anti-inflammatory properties, hence, balancing the actions of angiotensin II on the type 1 receptor.^14,15^ Concerns regarding both ACEI and ARBs have led to calls for discontinuation of these drugs both prophylactically and in those with suspected COVID-19.^15^ In contrast, using a large longitudinal population-based control group and a multi-centre cohort of COVID-19 cases, we have demonstrated that those on ARBs are significantly less likely (OR: 0.22 [95% CI 0.15-0.30]) to develop COVID-19. Potential benefit from ARBs in this context may be related to the effect of SARS-CoV-2 which once gaining entry through ACE2 into type II pneumocyte downregulates ACE2 leading to unabated angiotensin II induced organ injury. In a mouse model, SARS-CoV-1 induced lung injury could be limited by renin-angiotensin-aldosterone system blockade.^28^ Our analysis hasn’t shown ACEIs to have an effect similar to ARBs, consistent with the observations that ACEIs currently in clinical use do not directly affect ACE2 activity.^29^ We are not aware of any population based case-control studies thus far investigating risk factors increasing susceptibility to COVID-19, however, a systematic review^30^ found that ACEIs reduced the risk of pneumonia. In the latter study, the benefits of ACEI were substantially greater among Asian patients and these were attributed to a higher prevalence of ACE polymorphisms increasing the ACEI levels and kinin catabolism in this ethnic group.

Of 578 with COVID-19, 128 developed severe or critical form of the disease, higher proportion of patients (42.8%) with common form of the disease were on ARBs compared to severe/critical (35.1%) form of the disease. However, neither ARBs nor ACEI were significantly associated with the severity of COVID-19. A cohort study from Wuhan province, demonstrated lower (3.7%) 28-day all-cause mortality among 188 patients with COVID-19 receiving ARBs or ACEI in-hospital compared 9.8% among those who weren’t receiving these drugs.^31^ That study was not powered to evaluate the effect of ACEIs vs ARBs. In addition, lower proportion of patients with COVID-19 overall were on ARBs or ACEIs than expected^32,33^ raising the possibility that concerns regarding these drugs at the beginning of the epidemic may have resulted in change in medication in patients on admission, In-hospital use of ACEI has previously been shown to be associated with lower rate of ventilation and in-hospital mortality with viral pneumonias.^34^

Our case-control analysis showed that those taking CCBs had significantly increased risk [OR 1.73 (95% CI 1.2-2.3)] of manifesting symptoms of COVID-19. Two recent studies have shown consistent findings regarding the association between CCBs and increased risk of COVID-19 associated symptoms. ^35, 36^ The mechanisms underlying these findings are unclear as there is no evidence that either CCBs altering ACE2 expression. One study including 4792 hospitalised pneumonia (bacterial and viral) case, showed increased incidence of major cardiovascular events among those who were on CCBs, betablockers or corticosteroids.^37^ Impaired induction of anti-viral type 1 interferon response to a range of respiratory viruses has been well described in association with corticosteroids and may also apply in the context of COVID-19 explaining association of these drugs with the severity of infection in our cohort. ^38,39^

Hypertension was a strong risk factor for the development of the severe and critical forms of COVID-19 independently of antihypertensive therapy. This may be related to higher levels of endothelin in hypertensive patients and its effect on innate immune response. There is strong evidence that the innate immune response is key in the development of severe COVID-19.^40^ Recent data suggest that macrophages contribute to, and protect from, hypertension and that macrophage depletion augments the chronic hypertensive response to endothelin-1, (ET-1).^41^ High levels ET-1 are linked to lung damage in HIV infection.^42^ It is thus possible that arterial hypertension which is linked to higher levels of ET-1, more vasoconstriction, and lower blood flow, may be contributing to lower levels of macrophages (i.e. lower innate immune response) and potentially to build-up of fluid in the lungs (pulmonary oedema).

Our study has a number of strengths. The case-control design with cases from multiple hospitals and a large, prospective, population-based control group with 98% coverage of the local population has provided the power needed to identify independent risk factors including medication use to assess the risk factors for COVID-19 in this population. We have included consecutive cases from each of the secondary care hospitals to reflect the full range of case-mix seen in routine clinical practice. Case definition and assessment of severity of COVID-19 were adherent to national guidelines consistently across all the centers involved in this study.^16^

The limitations of our study include the retrospective nature of the COVID-19 cohort, which could lead to possible under-recording of some less common comorbidities and drug history, in particular related to use of glucocorticoids. Even though we included 578 cases, we may have modest power to evaluate potential risk factors with low frequency. In the large case series (n=1099) of COVID-19 patients,^3^ a number of co-morbidities such as cardiovascular (2.5%), cerebrovascular (1.4%) and chronic obstructive pulmonary diseases (1.1%) were infrequent and cancers, immunodeficiency and chronic kidney disease altogether formed 1.8% of the COVID-19 cases. Most patients with cardiovascular comorbidities qualify for angiotensin-converting enzyme inhibitor (ACEI) or angiotensin II receptor blocker (ARB) therapy. However, prescribing patterns of these drugs vary widely from China to the UK where ACEIs are much more commonly prescribed.^43^ Although with a much larger sample size this may prove to be significant if the effect is real, this compared poorly with other factors contributing to severity such hypertension (OR 2.8) or the use of immunosuppressants (OR 5.37). In addition, low case-fatality rate of 0.6% means that we were not able to assess risk factors associated with mortality. Another limitation is the lack of pre-pandemic data on glycemic control, since DPPIV inhibitors and insulin are usually prescribed to patients with poorer glycemic control than those receiving glucosidase inhibitors. Therefore, differences in severity seen between these medication categories could be reflecting differences in glycemic control and not necessarily an effect of the medication on disease progression.

Moreover, although all the medications included corresponded to prescriptions filled 3 or more times before diagnosis of COVID-19 we were unable to adjust for length of use of the medications and the current analysis did not adjust for multiple testing so some of the more modest associations are viewed with caution. Finally, some of the associations show strong statistical significance but, as is the case for CCBs, the odds ratios are < 2.0 and therefore they may not be of high clinical impact.

## Conclusion / Clinical relevance

We have identified higher BMI, diabetes and cardio/ cerebrovascular disease as independent risk factors for the development of COVID-19. Prior treatment with ARBs and diuretics was associated with reduced risk and CCBs with increased risk of developing COVID-19. Other antihypertensive drugs were not associated with increased risk of severe or critical form of the infection. So, we found no evidence to alter ARBs or ACEIs therapy in the context of the pandemic. We have also found first time, increased risk of COVID-19 in association with DPP4 inhibitors and insulin as reduced risk with glucosidase inhibitors. If replicated, these findings have clinical and potentially public health implications.

## Data Availability

Data used in the current analysis will be made available on request. The data has currently not been uploaded on a repository or online database.

## Author contributions

AMV, AV and GPA wrote the manuscript; HY, TC and GPA designed the research; HY, SW, LL, SY, HW, XT, JD, SJ, KH, FJ, SZ, NZ, YH, TC. performed the research; AV and AMV analyzed the data;.

## Collaborators

1. Department of Infectious Diseases, Hwamei Hospital, Ningbo No.2 Hospital, University of Chinese Academy of Sciences., Ningbo 315010, China;
2. NAFLD Research Center, Department of Hepatology, the First Affiliated Hospital of Wenzhou Medical University, Wenzhou 325000, China;
3. Department of Critical Care Medicine, Wenzhou Central Hospital, Wenzhou 325000, China;
4. Department of Infectious Diseases, Ruian People’s Hospital, Wenzhou 325200, China;
5. Department of Infectious Diseases, Shaoxing people’s Hospital, Shaoxing 312000, China;
6. Department of Infectious Diseases, Zhoushan Hospital, Zhoushan 316004, China;
7. Department of Infectious Diseases, Jiaxing First Hospital, 314000 Jiaxing, China;
8. Department of Infectious Diseases, Wenling First people’s Hospital, Taizhou 317500, China;
9. Department of Infectious Diseases, Hangzhou Xixi Hospital, Hangzhou 310023, China;
10. Department of Infectious Diseases, Taizhou Enze Medical Center (Group) Enze hospital, Taizhou 317000, China;
11. Department of Infectious Diseases, The First people’s Hospital of Yuhang District, Hangzhou 310006, China;
12. Department of Infectious Diseases, Ningbo Yinzhou people’s Hospital, Ningbo 315040, China;
13. Department of Infectious Diseases, Huzhou Central Hospital, Huzhou 313003, China;
14. Department of Infectious Diseases, Shulan (hangzhou) Hospital Affiliated to Zhejiang Shuren University Shulan International Medical College, Hangzhou 310004, China.

## References

1. WHO report-90. WHO, COVID-19 situation report-90. https://www.who.int/docs/default-source/coronaviruse/situation-reports/20200620-covid-19-sitrep-152.pdf?sfvrsn=83aff8ee_4. Accessed 21 June 2020

2. Wu Z. Characteristics of and Important Lessons From the Coronavirus Disease 2019 (COVID-19) Outbreak in China: Summary of a Report of 72314 Cases From the Chinese Center for Disease Control and Prevention. JAMA. 323 (13), 1239–1242 (2020).

3. Guan WJ.et al. Clinical Characteristics of Coronavirus Disease 2019 in China. N Engl J Med. 382, 1708–1720 (2020).

4. Shi Q. et al. Diabetic Patients with COVID-19, Characteristics and Outcome: A Two-Centre, Retrospective, Case Control Study. (2020); Available at http://dx.doi.org/10.2139/ssrn.3551369 (preprint)

5. Huang C.et al. Clinical features of patients infected with 2019 novel coronavirus in Wuhan, China. Lancet.395,497–506 (2020).

6. Yang X.et al. Clinical course and outcomes of critically ill patients with SARS-CoV-2 pneumonia in Wuhan, China: a single-centered, retrospective, observational study. Lancet Resp Med. 8(5), 475-481 (2020).

7. Li W.et al. Receptor and viral determinants of SARS-coronavirus adaptation to human ACE2. Nature. 24, 1634–43 (2005).

8. Hoffmann M.et al. SARS-CoV-2 Cell Entry Depends on ACE2 and TMPRSS2 and Is Blocked by a Clinically Proven Protease Inhibitor. Cell. 181, 271-280.e8 (2020).

9. Hamming I.et al. Tissue distribution of ACE2 protein, the functional receptor for SARS coronavirus. A first step in understanding SARS pathogenesis. J Pathol. 203, 631–7 (2004).

10. Dijkman R.et al. Replication-dependent downregulation of cellular angiotensin-converting enzyme 2 protein expression by human coronavirus NL63. J Gen Virol. 93, 1924–9 (2012).

11. Esler M, Esler D. Can angiotensin receptor-blocking drugs perhaps be harmful in the COVID-19 pandemic? J of Hypertension. 38, 781–2 (2020).

12. Fang L. et al. Are patients with hypertension and diabetes mellitus at increased risk for COVID-19 infection? Lancet Resp Med. 8(4), e21-2600(20)30116-8 (2020).

13. Aronson JK, Ferner RE. Drugs and the renin-angiotensin system in covid-19. BMJ. 369, m1313 (2020).

14. Li, G.et al. Antihypertensive treatment with ACEI/ARB of patients with COVID-19 complicated by hypertension. Hypertens Res. 43, 588–590 (2020).

15. Vaduganathan M.et al. Renin–Angiotensin–Aldosterone System Inhibitors in Patients with Covid-19. N Engl J Med. 382, 1653–1659 (2020).

16. Filardi T, Morano S.J COVID-19: is there a link between the course of infection and pharmacological agents in diabetes? J Endocrinol Invest. 3, 1–8 (2020).

17. National Health Commission of the people’s Republic of China. The Guidelines for the Diagnosis and Treatment of New Coronavirus Pneumonia (fifth edition). Available from: ww.nhc.gov.cn/yzygj/s7653p/202002/3b09b894ac9b4204a79db5b8912d4440.shtml

18. Lin H.et al. Using big data to improve cardiovascular care and outcomes in China: a protocol for the Chinese Electronic health Records Research in Yinzhou (CHERRY) Study. BMJ Open. 8, e019698 (2018).

19. Wang D.et al. Clinical characteristics of 138 hospitalized patients with 2019 novel coronavirus infected pneumonia in Wuhan, China. JAMA. 323(11), 1061–1069 (2018).

20. Vankadari N, Wilce JA. Emerging Wuhan (COVID-19) coronavirus: glycan shield and structure prediction of spike glycoprotein and its interaction with human CD26. Emerg Microbes Infect. 9(1), 601–604 (2020).

21. Zhao X.et al. Inhibition of Endoplasmic Reticulum-Resident Glucosidases Impairs Severe Acute Respiratory Syndrome Coronavirus and Human Coronavirus NL63 Spike Protein-Mediated Entry by Altering the Glycan Processing of Angiotensin I-Converting Enzyme 2. Antimicrobial Agents and Chemotherapy. 59(1), 206–216 (2014).

22. Bornstein S.et al. Endocrine and metabolic link to coronavirus infection. Nature Rev Edocrinology. 16, 297–298 (2020).

23. Williamson E. et al. Factors associated with COVID-19-related death using OpenSAFELY. Nature. (2020).

24. Bozkurt B. et al. HFSA/ACC/AHA Statement Addresses Concerns Re: Using RAAS Antagonists in COVID-19. American Heart Association Professional Heart Daily. March 19, (2020).

25. Phadke M, Saunik S. Rapid Response: Use of angiotensin receptor blockers such as Telmisartan, Losartsan in nCoV Wuhan Corona Virus infections – Novel mode of treatment. BMJ. 368, m406 (2020).

26. Ferrario CM.et al. Effect of Angiotensin-Converting Enzyme Inhibition and Angiotensin II Receptor Blockers on Cardiac Angiotensin-Converting Enzyme 2. Circulation. 111, 2605–2610 (2005).

27. Lu R.et al. Genomic characterisation and epidemiology of 2019 novel coronavirus: implications for virus origins and receptor binding. Lancet. 395(10224), 565–574 (2020).

28. Kuba K.et al. A crucial role of angiotensin converting enzyme 2 (ACE2) in SARS coronavirus-induced lung injury. Nat Med. 11, 875–9 (2005).

29. Rice GI, Thomas DA, Grant PJ, et al. Evaluation of angiotensin-converting enzyme (ACE), its homologue ACE2 and neprilysin in angiotensin peptide metabolism. Biochem J. 383, 45–51 (2004).

30. Caldeira D.et al. Risk of pneumonia associated with use of angiotensin converting enzyme inhibitors and angiotensin receptor blockers: systematic review and meta-analysis. BMJ. 345, e4260 (2012).

31. Zhang P.et al. Association of Inpatient Use of Angiotensin Converting Enzyme Inhibitors and Angiotensin II Receptor Blockers with Mortality Among Patients With Hypertension Hospitalized With COVID-19. Circulation Res. 126, 1671–1681 (2020).

32. Wang Z. et al. Status of hypertension in China: results from the China Hypertension Survey, 2012-2015.Circulation. 137, 2344–56 (2018).

33. Lu J.et al. Prevalence, awareness, treatment, and control of hypertension in China: data from 1・ 7 million adults in a population-based screening study (China PEACE Million Persons Project). Lancet. 390, 2549–58 (2017).

34. Henry C.et al. Impact of angiotensin-converting enzyme inhibitors and statins on viral pneumonia. Baylor Univ Med Cent Proc. 31, 419–423 (2018).

35. Reynolds HR.et al. Renin–angiotensin–aldosterone system inhibitors and risk of Covid-19. N Engl J Med. 382, 2441–2448 (2020).

36. Mancia G.et al. Guidelines for the management of hypertension and target organ damage. J of Hypertension. 31 (12), 2464-2465 (2013).

37. Steiner GS. et al. Bacterial pneumonia compared to viral pneumonia is associated with a higher risk of future major adverse cardiovascular events. Presented at: American Heart Association Scientific Sessions 2018; November 10-12, 2018; Chicago, IL. Available at: https://www.abstractsonline.com/pp8/#!/4682/presentation/51526

38. Singanayagam, A.et al. Corticosteroid suppression of antiviral immunity increases bacterial loads and mucus production in COPD exacerbations. Nat Commun. 9, 2229 (2018).

39. Thomas, B. et al. Glucocorticosteroids enhance replication of respiratory viruses: effect of adjuvant interferon. Sci Rep. 4, 7176 (2015).

40. Shi Y.et al. COVID-19 infection: the perspective of immune response. Cell Death & Diff. 27, 1451–1454 (2020).

41. Czopek A.et al. A novel role for myeloid endothelin-B receptors in hypertension. Eur Heart J. 40(9), 768–784 (2020).

42. Head BM.et al. Inflammatory mediators and lung abnormalities in HIV: A systematic review. Plos One. 14(12), e0226347 (2019).

43. Mahmoudpour HS.et al. Prescription patterns of angiotensin-converting enzyme inhibitors for various indications: A UK population-based study. Br J Clin Pharmacol. 84(10), 2365–2372 (2018).

